# Did “long COVID” increase road deaths in the U.S.?

**DOI:** 10.1101/2023.10.11.23296868

**Authors:** Leon S. Robertson

**Author notes:** Disclosure: This research was not funded. The author has no financial or other interests that would be affected by publication. Data availability: Data sources are publically available at the websites cited.

## Abstract

**Objective:** To examine data on COVID-19 disease associated with a 10 percent increase in U.S. road deaths from 2020 to 2021 that raises the question of the potential effect of pandemic stress and neurological damage from COVID-19 disease.

**Methods:** Poisson regression was used to estimate the association of recent COVID-19 cases, accumulated cases, maximum temperatures, truck registrations, and gasoline prices with road deaths monthly among U.S. states in 2021. Using the regression coefficients, changes in each risk factor from 2020 to 2021 were used to calculate expected deaths in 2021 if each factor had remained the same as in 2020.

**Results:** Corrected for the other risk factors, road deaths were associated with accumulated COVID-19 cases but not cases in the previous month. More than 20,700 road deaths were associated with the changes in accumulated COVID-19 cases but were substantially offset by about 19,100 less-than-expected deaths associated with increased gasoline prices.

**Conclusions:** While more research is needed, the data are sufficient to warn people with “long COVID” to minimize road use.

**What is already known about this topic:** Previous short-term fluctuations in road deaths are related to changes in temperature, fuel prices, and truck registrations.

**What this study adds:** Corrected for other risk factors, the monthly changes in road deaths from 2020 to 2021 in U.S. states were associated with cumulative COVID-19 cases.

**How this study might affect research, practice, or policy:** Studies are needed to distinguish the potential relative effects of neurological damage as well as the stress of coping with the pandemic on driving, walking, and bicyclist behavior. Warning people with “long covid” about road risk is warranted.

## Introduction

While many countries experienced a reduction in road deaths during the first year of the COVID-19 pandemic^1^, deaths increased 6.9 percent from 2019 to 2020 in the U.S. In 2021 road deaths in the U.S. increased 10.5 percent above those experienced in 2020^2^. Analysis of the 2020 increase among U.S. states found most of it correlated to changes in temperatures, truck registrations, and fuel prices. The increase month by month was not significantly correlated with COVID-19 cases in the prior month among U.S. states, controlling statistically for the three mentioned risk factors^3^. Fluctuations in monthly state road deaths have been related consistently to changes in temperatures, truck registrations, and fuel prices for decades^4^. Fatal vehicle crashes occur disproportionately at night and on weekends when traffic is lighter. Temperature and gas prices are proxy variables for discretionary travel. Trucks are disproportionately involved in fatal crashes.

In 2021 The U.S. average maximum temperature averaged monthly among the states, changed little from the average in 2020. Truck registrations increased 4.4 percent but fuel prices rose 38 percent. The latter should have more than offset the effect of the increase in truck registrations on the road death rate per population. The possible role of COVID-19 as a risk factor for the increased road deaths in 2021 deserves a closer look.

The neurological changes in people with COVID-19 disease include the potential for an increase in road deaths. A wide variety of neurological symptoms occur in the aftermath of infection of many people ^5-7^ and changes in the brains of those infected have been observed^8^. Rather than an association of recent COVID-19 cases and road deaths, a “long covid” effect would be associated with accumulated cases. The purpose of this paper is to report a test of the hypothesis that differences in road deaths among U.S. states in 2021 are associated with accumulated COVID-19 cases, controlling statistically for the other mentioned risk factors.

## Materials and Methods

Data on the monthly average of daily high temperature^9^, truck registrations,^10^ and gasoline prices^11^ were matched to COVID-19 cases, accumulated cases^12^, and road deaths^13^ for each U.S. state during 2020-2021. COVID-19 cases and accumulated cases were the numbers at the beginning of the month while the others were matched in the same month as the road deaths. Temperature data for each weather station in a given state and month were averaged. Truck registrations are reported annually and are prorated by month. Monthly gasoline prices are national averages. They vary somewhat among and within states.

The 2021 data were fitted to a Poisson regression model of the form:

State monthly road deaths =

b1 x (average high temperature) + b2 x (gas price) +

b3 x (truck registrations/population) +

b4 x (COVID-19 previous month cases/population x100) +

b5 x (accumulated COVID-19 cases/population*100) +

b6 x (March=1, else 0) +

b7 x (April=1, else 0) +

b8 x (May=1, else 0).

Log(population) was included as an offset variable to correct for differences among the states. The state population data are from the 2020 census^14^. Binary variables for March, April, and May are included because those were the months of the shutdowns in most U.S. states and were included in the model that found no correlation between road deaths to monthly COVID cases^3^.

To assess the number of road deaths associated with changes in the predictor variables, the regression coefficients were used to estimate the number of deaths in each month in each state that would have occurred if each variable had remained at the 2020 observation. The differences between those estimates and the number that occurred in 2021 were summed across the states. No patients or public were involved in the design or execution of this study.

## Results

The percent increase in road deaths from 2020 to 2021 was substantially the result of the increase in March through May as indicated in Figure 1. March-May 2020 was the period of stay-at-home orders in most U.S. states in 2020 and motor vehicle use declined as a large segment of the population complied^2^.

**Figure 1.**
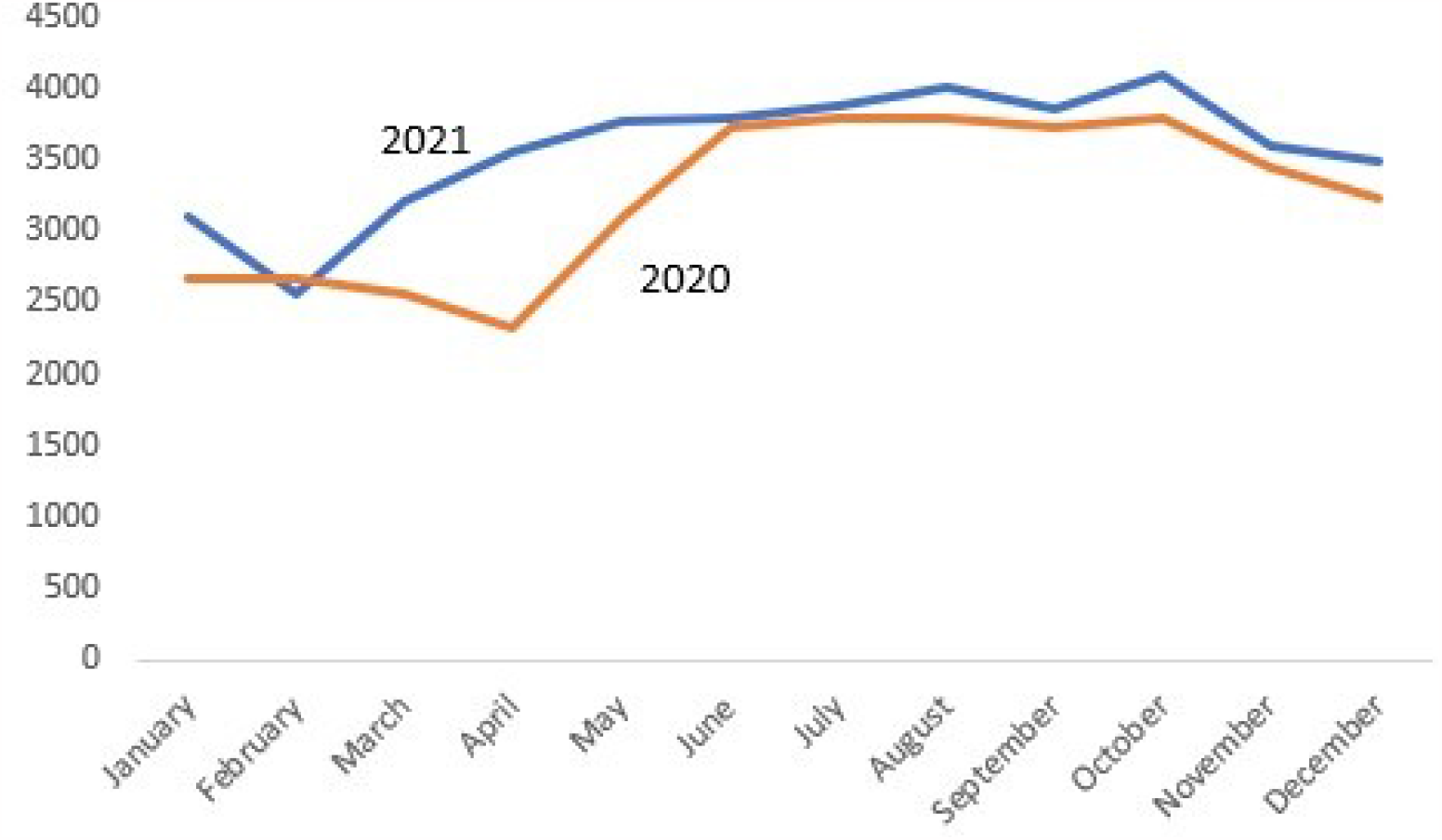
Monthly Road Deaths in the U.S., 2020-2021.

The least squares correlation between the deaths predicted by the regression model and actual deaths by month among the states in 2021 was 0.91, a good fit. Table 1 presents the odds ratios based on the regression coefficients and 95 percent confidence intervals. Increased road deaths are related to higher temperatures, lower gas prices, and more trucks per population as found in previous years. There is no significant correlation to COVID-19 cases in the previous month but cumulative cases predict additional road deaths.

**Table 1.** Odds ratios based on Poisson regression predictors of road deaths in U.S. states by month, 2021.

| Variable | Odds Ratio | 95 % C.I. |
| --- | --- | --- |
| Temperature | 1.008 | 1.007, 1.009 |
| Gas price | .665 | .626, .706 |
| Trucks/population | 1.994 | 1.821, 2.184 |
| COVID-19 cases/100 population | 1.002 | .985, 1.020 |
| Cumulative COVID-19 cases/100 population | 1.074 | 1.069, 1.079 |
| March | 1.041 | 1.001, 1.082 |
| April | 1.070 | 1.031, 1.111 |
| May | 1.076 | 1.043, 1.114 |

**Table 2.**
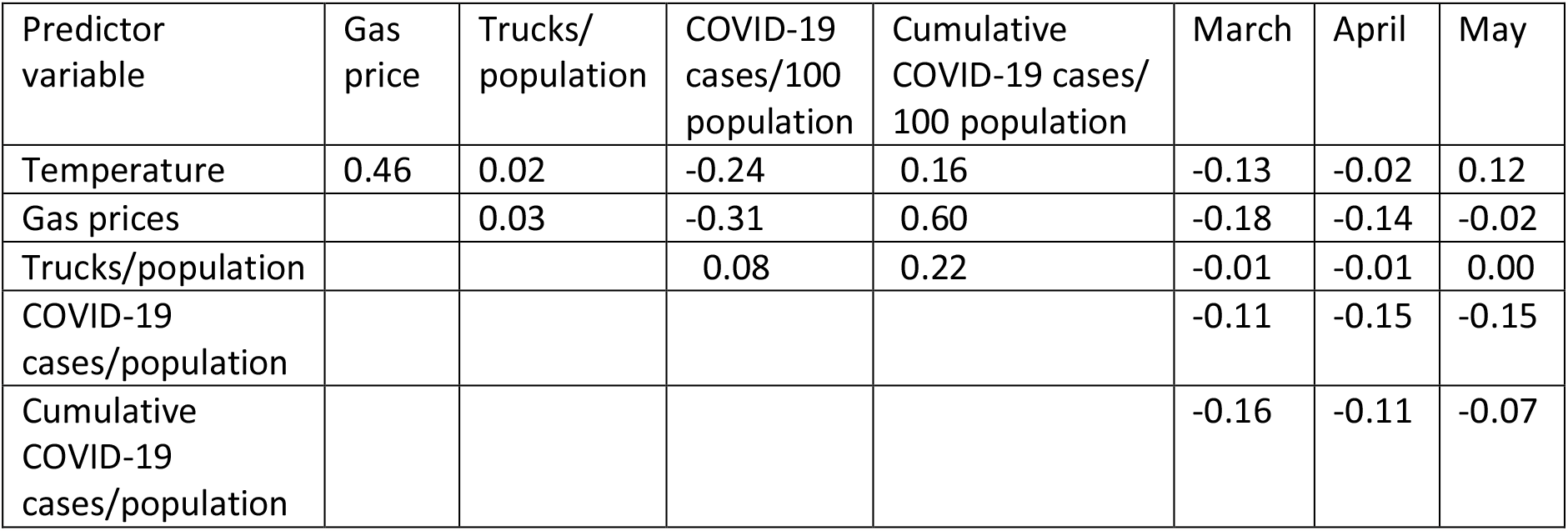
Least squares correlations among predictor variables.

| Predictor variable | Gas price | Trucks/population | COVID-19 cases/100 population | Cumulative COVID-19 cases/100 population | March | April | May |
| --- | --- | --- | --- | --- | --- | --- | --- |
| Temperature | 0.46 | 0.02 | -0.24 | 0.16 | -0.13 | -0.02 | 0.12 |
| Gas prices |  | 0.03 | -0.31 | 0.60 | -0.18 | -0.14 | -0.02 |
| Trucks/population |  |  | 0.08 | 0.22 | -0.01 | -0.01 | 0.00 |
| COVID-19 cases/population |  |  |  |  | -0.11 | -0.15 | -0.15 |
| Cumulative COVID-19 cases/population |  |  |  |  | -0.16 | -0.11 | -0.07 |

The correlations among the predictor variables were examined for potential distortion of the coefficients by collinearity. Most of the correlations are remarkably low and none are high enough to be worrisome. Gas prices increased more in states that had a higher rate of accumulated cases per population but the correlation is not strong enough to distort the regression coefficients substantially.

The estimated difference in road deaths in 2021 if each risk factor had remained the same while the others changed is shown in Table 3. Had gas prices remained as low as in 2020, the model predicts that 19,078 additional deaths would have occurred in 2021. If the correlation of road deaths with accumulated COVID-19 cases is causally linked and if COVID-19 had been contained, some 20,698 road deaths in 2021 would have been prevented.

**Table 3.** Differences in expected 2021 road deaths if each risk factor remained the same as in 2020.

| Variable | Difference |
| --- | --- |
| Temperature | 77 |
| Gas price | -19078 |
| Trucks/Population | 572 |
| Cumulative COVID-19 cases/Population | 20698 |
| Total | 2391 |

## Discussion

These results suggest that the road deaths related to cumulative COVID-19 cases were largely offset by increased fuel costs in 2021. If fuel costs had remained low, the increase in total deaths would have far exceeded the aggregate 10.5 percent increase. But are the 20,000 plus road deaths related to cumulative cases of COVID-19 the result of neurological damage by the SARS-CoV-2 virus?

While a “long-COVID” effect is plausible given the symptoms, this question cannot be answered by aggregated data. The lack of association of road deaths with the previous months’ cases suggests that the problem is not the short-term stress of managing life when the virus is a more immediate threat. That does not rule out stress and distractions exacerbated by dealing with prolonged surges of infections. While the symptoms called “long COVID” can persist for months^15^, if brain damage occurs, there could be subtle changes in behavior not identified as symptoms far longer, even permanently.

Case-control studies comparing persons in crashes while driving or struck by vehicles with others at the scene at the same time of day and day of the week would be more definitive. Questions that should be included in such studies are: Have these persons been diagnosed with COVID-19 in the past? If so, how long ago? Did they have symptomatic neurological problems and, if so, how recently? Did they lose a family member or friend to COVID-19 infection and, if so, how long ago? Are they driving, walking, or bicycling at the crash scene as part of usual behavior before the pandemic or for some reason related to COVID-19 avoidance or treatment? What drugs or other therapy, if any, are they using to cope with the pandemic in general or “long-COVID” symptoms in particular?

In the absence of such studies, the hypothesis has sufficient plausibility to warrant cautioning people who have been infected with SARS-CoV-2 and its mutated variants to avoid driving if possible or be extremely cautious when on the road as a driver, pedestrian or bicyclist..

## Data Availability

9. National Centers for Environmental Information. Climate data online. National Oceanic and Atmospheric Administration. March 2023: https://www.ncdc.noaa.gov/cdo-web/
10. Federal Highway Administration. State motor-vehicle registrations. Highway Statistics 2021. March 2023. https://www.fhwa.dot.gov/policyinformation/statistics/2021/
11. U.S. Energy Information Administration. Petroleum and other liquids. March 10, 2022. https://www.eia.gov/dnav/pet/hist/LeafHandler.ashx?n=pet&s=emm_epm0_pte_nus_dpg&f=m
12. U.S. Centers for Disease Control and Prevention. United States COVID-19 cases and deaths by state over time. March 10, 2023. https://data.cdc.gov/Case-Surveillance/United-States-COVID-19-Cases-and-Deaths-by-State-o/9mfq-cb36
13. National Highway Traffic Safety Administration. Fatality Analysis Reporting System. May 2022. https://www.nhtsa.gov/research-data/fatality-analysis-reporting-system-fars
14. U.S. Census Bureau. National population totals and components of change: 2020-2021. March 2023. https://www.census.gov/data/datasets/time-series/demo/popest/2020s-national-total.html

## Notes

### Competing Interest Statement

The authors have declared no competing interest.

### Funding Statement

This study did not receive any funding.

### Author Declarations

The study used (or will use) ONLY openly available human data that were originally located at:9.National Centers for Environmental Information. Climate data online. National Oceanic and Atmospheric Administration. March 2023: https://www.ncdc.noaa.gov/cdo-web/ 10.Federal Highway Administration. State motor-vehicle registrations. Highway Statistics 2021. March 2023. https://www.fhwa.dot.gov/policyinformation/statistics/2021/ 11.U.S. Energy Information Administration. Petroleum and other liquids. March 10, 2022. https://www.eia.gov/dnav/pet/hist/LeafHandler.ashx?n=pet&s=emm_epm0_pte_nus_dpg&f=m 12.U.S. Centers for Disease Control and Prevention. United States COVID-19 cases and deaths by state over time. March 10, 2023. https://data.cdc.gov/Case-Surveillance/United-States-COVID-19-Cases-and-Deaths-by-State-o/9mfq-cb36 13.National Highway Traffic Safety Administration. Fatality Analysis Reporting System. May 2022. https://www.nhtsa.gov/research-data/fatality-analysis-reporting-system-fars 14.U.S. Census Bureau. National population totals and components of change: 2020-2021. March 2023. https://www.census.gov/data/datasets/time-series/demo/popest/2020s-national-total.html

